# Safety and Efficacy of Therapeutic Hypothermia in Acute Traumatic Spinal Cord Injury: A Systematic Review and Meta-Analysis of human evidence-based studies

**DOI:** 10.1101/2025.10.24.25338722

**Authors:** Farzan Fahim, Mahdi Mehmandoost, Pouya Karami Dehkordi, Ali Khorram, Shahriar Heshmaty, Fatemeh Zolfaghari, Mobina Ghamarpour, Sayeh Oveisi, Amir Saied Seddighi, Alireza Zali

**Affiliations:** Neurosurgery Resident, Shohada Tajrish Hospital, Shahid Beheshti University of Medical Sciences, Tehran, Iran; Functional Neurosurgery Research Center, Research Institute of Functional Neurosurgery, Shohada Tajrish Comprehensive Neurosurgical Center of Excellence, Shahid Beheshti University of Medical Sciences, Tehran, Iran; Student Research Committee, School of Medicine, Shahid Beheshti University of Medical Sciences, Tehran, Iran; Student Research Committee, School of Medicine, Zanjan University of Medical Sciences, Zanjan, Iran; Student Research Committee, School of Medicine, Hormozgan University of Medical Sciences, Bandar Abbas, Iran; Medical student at Isfahan University of Medical Science, Isfahan, Iran; Medical Student at Shahid Beheshti University of Medical Sciences, Tehran, Iran; Medical Student at Kurdistan University of Medical Science, Student Research Committee, Sanandaj, Iran; Microbiology Student, Tehran Azad University of Medical Science, Tehran, Iran; Professor of Neurosurgery, Shohada Tajrish Hospital, Shahid Beheshti University of Medical Science, Tehran, Iran

**Keywords:** traumatic spinal cord injury, therapeutic hypothermia, cooling, function recovery, meta-analysis, neurologic recovery, critical care

## Abstract

**Background:** Therapeutic hypothermia as a neuroprotective strategy can reduce secondary injury after acute traumatic spinal cord injury (SCI). Although promising in animal models, its clinical effectiveness and safety remain uncertain. We conducted a systematic review and meta-analysis on human studies to evaluate the impact of systemic or local hypothermia on neurological outcomes, mortality, and length intensive care hospitalization.

**Methods:** Systematic and comprehensive search in PubMed, Scopus, Web of Science, Embase, and Cochrane Trails with no limitation of time and language performed. Following PRISMA 2020 guidelines, two independent reviewers screened records across four databases up to 20 September 2025. Eligibility was determined using PICOS criteria: adults with acute traumatic SCI (P), receiving any hypothermic protocol (I), compared with standard normothermic management (C), with reported functional, sensory, or survival outcomes (O), and original human clinical designs (S). Animal experiments, case reports, and conference abstracts without full texts were excluded. Data extraction and risk-of-bias evaluation were performed independently using the Joanna Briggs Institute (JBI) checklist. Pooled relative risks (RR) or mean differences (MD) were calculated with random-effects models (DerSimonian-Laird) using RevMan 4.5.1

**Results:** From 612 initial records, six human studies met inclusion criteria: three systemic hypothermia trial and one local extradural protocol, plus two supportive cohorts providing timing and assessment data. Intervention methods included surface, and endovascular techniques maintaining body temperature at 32-34 Celsius for 24-72 hours, initiated between 1.6 -70 hours post-injury. pooled analyses showed Mortality with RR = 0.57 (95 % CI 0.05 -5.88), indicating no significant difference but a trend favoring hypothermia. AIS grade improvement with RR = 2.96 (95 % CI 0.01-939.31); direction toward neurological benefit though statistically imprecise. ICU length of stay with MD = -1.27 days (95 % CI -2.46 to -0.07), suggesting shorter intensive care duration. complications included pneumonia, hypotension, and bradycardia, with no hypothermia-related deaths were reported. Early initiation (< 6 h) was consistently linked with superior functional improvement. Overall methodological quality was low-to-moderate; none of the studies were randomized.

**Conclusions:** Results from six human studies reveal that therapeutic hypothermia may be feasible, safe, and effective as an intervention in acute traumatic SCI. While current evidence cannot yet demonstrate mortality benefit, the observed results were directed toward neurological improvement, especially when therapy is initiated early and systemically, supports ongoing investigation. Until large randomized data are available, hypothermia should be regarded as an experimental yet promising neuroprotective additional therapeutic option and complementing early decompression.

## Introduction

Traumatic Spinal cord injury (tSCI) is a major burden of global public health because of its high mortality, long-term morbidity, and marked socioeconomic consequences. Recent global estimates reported approximately 0.9 million new cases of tSCI in 2019, with more than 20.6 million individuals living with SCI worldwide, which involve more than 6.2 million years lived with disability (YLDs)[1]. The incidence of traumatic SCI remains considerable suggesting an average of near 26.5 cases per million population annually[2]. Cervical injuries, most common among these injured patients, are associated with respiratory disabilities, higher mortality, and more severe neurological sequelae [3].

Despite advances in acute trauma care, mortality remains significantly high among tSCI patients compared to the general population. Especially tSCI in older adults faces them into poor survival prospects; for example, based on data from US population, one-year mortality in elderly patients with cervical fracture-SCI is several-fold higher than in younger cohorts[4].Survivors experience severe and chronic morbidities, including motor paralysis, bladder and bowel dysfunction, chronic pain, respiratory complications, infections, and pressure ulcers, which together decrease quality of life and reduce life expectancy by 10–15 years on average[5].Thoracic and lumbar injuries show distinct morbidity profiles, but, overall mortality is lower than in cervical SCI population. Thoracic SCI often results in paraplegia, autonomic dysfunction including bladder and bowel dysfunction, loss of trunk control, and suppressed cardiovascular responses; high thoracic lesions may still affect sympathetic chain leading to hypotension. Lumbar injuries frequently impair lower limb motor and sensory function, gait, and cause chronic pain, also cause similar autonomic disturbances which is less severe than higher levels injuries. The incidence and risk of secondary complications such as pressure ulcers, urinary tract infections, respiratory infections (especially in high thoracic injuries), deep vein thrombosis, and osteoporosis remain high. recovery rates are lower: a study showed that thoracic SCI patients have significantly reduced likelihood of considerable neurologic recovery compared to cervical lesions [6].staying hospital days and rehabilitation periods are often higher ; for example, in a large recent review, thoracic insults showed about 35% of all traumatic SCI cases, with patients showing longer ICU and post ICU care needs and staying and higher rates of readmission in hospital for dermatologic and genitourinary complications[7].

The economic impact of tSCI is profound. Some Systematic reviews have estimated direct costs of lifetime ranging from US$0.7 million to more than US$2.5 million per patient, which depend on age and severity of injury[8, 9]. First-year hospitalization and rehabilitation costs alone is highert than US$330,000 in high-income countries, while indirect costs related to lost productivity, long-term caregiving, and reduced social participation further amplify the economic burden[9]. These issues emphasize the necessity of effective interventions to eradicate secondary injury insults and improving functional outcomes.

Surgical decompression and stabilization main intervention for acute tSCI management, aiming to relieve cord compression, restore alignment, and create an optimized microenvironment for chance of recovery. Recent systematic reviews and meta-analyses have demonstrated that early decompression, particularly within 24 hours, is associated with improved motor outcomes, reduced complications, and shorter hospital stay[10].non-surgical interventions, such as the selective use of anti-inflammatory drugs, adjusted to surgery; however, high-dose methylprednisolone has shown limited neurological benefit and increased complication risks in recent randomized and cohort studies, making controversial in the setting of management [11, 12]. Therapeutic hypothermia has also been investigated as a perioperative adjunct to decompression, with pilot studies supporting feasibility and short-term safety, of-course larger randomized trials are still required to examine or establish its efficacy[13]. Regenerative approaches, including biomaterial scaffolds and cell-based therapies, are increasingly combined with surgical stabilization to modulate scarring, and trigger axonal regenerating, with several early-phase clinical trials reporting modest but hopeful results[14]. Furthermore, neuromodulation techniques such as epidural or transcutaneous spinal cord stimulation are often introduced during the rehabilitation stage after surgical fixation [15]. Altogether, decompressive surgery plays an essential role in management of acute tSCI, multimodal parallel therapies including pharmacological, hypothermic, regenerative, and neuromodulatory are synergistic strategies to maximize recovery from injuries.

For structural repair and regeneration, biomaterial scaffolds and extracellular matrix modifying strategies are being developed and tested in combination with cellular therapies. Cell transplantation using neural precursor cells, mesenchymal stromal cells (MSCs), and other progenitor cell types has demonstrated safety in many clinical studies and occasional modest functional gains, but trials are heterogeneous with respect to cell type, dose, timing, delivery route, and outcome measures, limiting definitive claims of efficacy[16]. Neuromodulation approaches, including epidural spinal cord stimulation and noninvasive transcutaneous stimulation, when paired with task-specific rehabilitation, have produced striking functional improvements in selected patients (including restoration of volitional standing and assisted walking in chronic cases), and they are an important and rapidly advancing therapeutic avenue[17]. Contemporary rehabilitation also integrates intensive task-specific training, robotic/exoskeleton assistance, and neuroprosthetic technologies to maximize recovery and independence.

Currently, no curative therapies exist to restore spinal cord function. at present, therapeutic strategies target the secondary insults, which start during hours to weeks following the primary mechanical trauma. Secondary injury involves a complex interaction of ischemic injury, glutamate excitotoxicity cascades, oxidative stress, ionic imbalance, mitochondrial dysfunction, and neuroinflammation, ultimately leading to neuronal and glial cell death, demyelination, and cavitation[18]. Vascular disruption and hemorrhage trigger early ischemia and hypoxia, while glutamate release and calcium overload initiate widespread excitotoxic damage[19]. Concurrently, production of reactive oxygen species and lipid peroxidation contribute to oxidative stress, damaging membranes and organelles[20]. Infiltration of neutrophils and macrophages, neuroinflammation done by microglial activation, and release of pro-inflammatory cytokines such as TNF-α and IL-1β can lead to exacerbatation of tissue loss and creation of a non-permissive environment for axonal regeneration [21]. These processes not only worsen the initial neurological deficit but also limit the potential for spontaneous repair. Consequently, most emerging therapeutic approaches including surgical decompression, pharmacological neuroprotection, hypothermia, and regenerative strategies are intervened to prevent secondary injury cascades and to preserve the chance for recovery of function [20].

Evidence supporting the efficacy of hypothermia in improving neurological outcomes remains heterogeneous [22,23]. Initial management focuses on rapid cervical spine immobilization, cardiorespiratory stabilization, and early spinal decompression and fixation when indicated [24]. Therapeutic hypothermia (local or systemic) has been investigated as a potential neuroprotective intervention because of its ability to reduce metabolic demand, inflammation, and apoptotic pathways. A recent meta-analysis reported that many numbers of treated patients with hypothermia as an adjusted therapy, experienced neurological improvement, which suggest a possible benefit; however, the underlying studies were small, methodologically diverse, and frequently uncontrolled [22]. preclinical and translational studies support the biologic hypothesis for hypothermia as a hopeful strategy but also highlight important limitations. Animal experiments suggest that its protective effects depend on factors such as timing, duration, and degree of cooling, and benefits may diminish with suboptimal protocols or after rewarming[25]. Clinically, prospective case series and pilot randomized studies have shown that systemic hypothermia is feasible and generally safe; however, reported effects on neurological recovery remain inconsistent. some studies describe motor or sensory improvements, while others find only minimal or no benefit[22, 25]. These discrepancies may reflect differences in patient selection, cooling methods, and outcome assessment. Overally, therapeutic hypothermia should be regarded as a promising but not yet established intervention for acute tSCI.

A systematic review is essential to clarify hypothermia intervention in clinical settings, because of high global burden of SCI, the prominent rate of mortality and morbidity associated with tSCI, and the uncertain evidence supporting the role of hypothermia as adjust strategy. Previous systematic reviews and meta-analyses have generally adopted a broad perspective, pooling highly heterogeneous interventions including pharmacological agents, surgical decompression, stem cell therapies, biomaterials, and rehabilitation protocols. While such reviews have been valuable for mapping the overall therapeutic landscape, the heterogeneity of interventions has limited the ability to draw conclusions specific to any single treatment, particularly therapeutic hypothermia. Earlier reviews have often combined cervical, thoracic, and lumbar injuries despite their distinct clinical and prognostic features, and in some cases have merged experimental animal data with small or uncontrolled human studies, reducing the strength of clinical inference. Moreover, many of these reviews predate recent feasibility trials and pilot randomized studies published after 2020, which provide important new insights. Therefore, a systematic review and meta-analysis focusing exclusively on therapeutic hypothermia in acute traumatic SCI is warranted to synthesize the latest clinical evidence, resolve discrepancies across studies, and provide a clearer basis for clinical decision-making and future trial design.

## Method

### Protocol and registration

This study was conducted in alignment with Preferred Reporting Items for Systematic Reviews and Meta-analyses (PRISMA) guidelines, version, 2020, and was registered in PROSPERO database (PROSPERO ID: CRD420251173039).

### Information Sources and search strategies

We covered a various database, PubMed, Scopus, Web of Science, Cochrane and Embase. Limitation of time and language were not applied. If any other than English studies were found in our search strategy, we planned to translate them into English before starting screening. The search strategy was based MeSH entry terms in MeSH database and keywords related to our title and PICO. The core search strategy words were Spinal Cord Injury, Hypothermia, Function Recovery, Motor and Sensory Recovery and Anti-inflammation. The complete search strategy syntax was illustrated in **supplementary 1**.

Each record of databases was extracted and after collecting all of them, they were entered For example, our search syntax of Scopus is:

TITLE-ABS-KEY (“Spinal Cord Injury” OR “SCI” OR “acute spinal cord trauma” OR “spinal

cord trauma”)

AND

(

TITLE-ABS-KEY (“Hypothermia, Induced” OR “therapeutic hypothermia” OR “systemic

hypothermia” OR “local hypothermia” OR “cooling therapy” OR “targeted temperature

management” OR “regional hypothermia”)

OR

INDEXTERMS (“hypothermia”)

)

AND

(

TITLE-ABS-KEY (“Recovery of Function” OR “neurological recovery” OR “motor recovery”

OR “sensory recovery” OR “neuroprotection” OR “secondary injury” OR “inflammation” OR

“decreasing second insult” OR “anti inflammatory”)

OR

INDEXTERMS (“recovery” OR “neuroprotection”)

)

AND

(

TITLE-ABS-KEY (“acute” OR “early phase” OR “post-injury” OR “subacute”)

)

Also, our full search strategy is attached as supplementary 1.

### Study selection

After defining the PICO of study and retrieving studies from all databases, Rayyan’s 2025, which is an online screening platform for systematic and meta-analysis, is based on its AI structure. Initially 531 studies were entered, and after removing the duplicates, 303 studies were left.

The screening process was conducted in two phases. In the first phase, two reviewers, P.K.D and S.A., independently performed titles and abstracts screening. each reviewer independently fills the excluded sheets involving these columns: study ID and DOI, reason of exclusion, which attached as **supplementary 2a and 2b**. Relative and original studies, except case reports, were included to be assessed for their eligibility in the second phase. The third reviewer (MM) solved the conflicts between the two reviewers. In the second phase, the full-text of each included study was assessed, two independent reviewer P.K.D and A.KH assessed each study by considering the PICO which was defining the participants with SCI injury who were under a hypothermia treatment method for finding out the outcomes of the surgeries and rehabilitation. The reviewers separately modified two Excell 2016 sheets, inclusion and exclusion studies, which the inclusion sheet details of study consisted title, first author, year of study, PICO and study design.

The exclusion sheet was consisted (Supplementary 2a and 2b), Title, first author, year of study and the reason for exclusion.

Animal studies weren’t excluded in the initial screening and they were also assessed in the second screen process. As same as the initial screening, the third author (MM) resolved the conflicts between the two reviewers, which could be any disagreement in defining the type of hypothermia or the type of spinal cord injury. Finally, based on the limited novelty for meta-analysis the animal studies were excluded, they were 6 human studies which exactly met our PICO and eligibility criterias were included for further analysis.

### Inclusion and exclusion criteria

We defined our **inclusion criteria** based on PICOS (participants, interventions, comparisons, outcomes, and study design).

1. Participants (P) Participants after any spinal cord injury which was in an acute phase
2. Intervention (I) Hypothermia and cooling procedure
3. Comparison(C) Patients with SCI who didn’t receive hypothermia as therapeutic intervention Outcome (O) Improving SCI patients, motor function, sensory function, improving quality of life and, rehabilitation.
4. Type of study All human studies such as cohort, case series, clinical trials were included.

### Exclusion criteria

Non-original studies were excluded. Case reports and Conferences studies which were without full-text were excluded.

Studies which didn’t investigated the hypothermia as an intervention or their injury side wasn’t localized in the spinal cord or the injury wasn’t occurred in an acute phase, were excluded as in an exclusion Excell sheet. Finally, animal studies were excluded.

#### Data extraction

Data extraction designed by FF, and was conducted independently by two reviewers (FZ and AK) (**Supplementary 3**). Each reviewer conducted the extraction from included data which was based on our inclusion criteria. The third reviewer (FF) resolved the conflicts between these researchers. Moreover, a structured data extraction sheet was conducted which included the following columns: Title, Authors and year of publication, DOI, funding sources and conflict of interest, country, study design, center type, time period of data collection, inclusion criteria, exclusion criteria, follow-up duration, ethical approval, age, male sex, mechanism of injury, NLI (Neurological level of injury), time from injury to intervention initiation, time from Injury to Enrollment, AIS/SCIM/FIM/WISCI/mRS/Other, AIS(A,B,C,D,E), AIS Motor Score (0-5), ASIA Sensory Score, SCIM, FIM, mRS, type of hypothermia, timing of initiation post-injury, cooling initiation setting ((ER, ICU, Intraoperative)), target temperature, duration of hypothermia (Total Cooling Period), Time to Reach target temperature (Hours), rewarming Protocol(Rate/Hour), active warning Post-rewarming (Yes,No,Method), temperature monitoring method(Esophageal, Bladder, Intravascular), use of sedation or neuromuscular blockage during cooling(Yes/No), concurrent treatment, type of surgery, steroid type (Dosage, Duration of Use), improvement in AIS grade, improvement in AIS grade, improvement in AIS grade, change in ASIS motor score, mortality rate, change in ASIS sensory score, change in SCIM score, change in FIM score, change in quality of life score, length of ICU stay, length of ICU stay, safety and adverse outcomes, timing of outcomes assessments, concomitant injury, static adjustment and adverse event.

### Risk of Bias assessment

We assessed the method quality using Joanna Briggs Institute’s (JBI) Critical appraisal Checklist, based on the study design. Two independent reviewers (MB and AK) who were not participate in primary included studies, performed the ROB assessment. **(Table 1.) (Supplementary 4a and 4b).**

**Table 1.** The studies risk of bias assessment using JBI 2020 checklists.

|  | Q1 | Q2 | Q3 | Q4 | Q5 | Q6 | Q7 | Q8 | Q9 | Q10 | Q11 | Overall |
| --- | --- | --- | --- | --- | --- | --- | --- | --- | --- | --- | --- | --- |
| **Batchelor2023** | ⊗ | ✔ | ✔ | ✔ | ✔ | unclear | ✔ | ✔ | unclear | ⊗ | ✔ | moderate |
| **battistuzzo2016** | ✔ | ✔ | ✔ | ✔ | ✔ | ✔ | ✔ | ✔ | ⊗ | ✔ | ✔ | Low |
| gallagher2020 | ✓ | ✓ | ✓ | ✓ | ✓ | ✓ | ✓ | ✓ | ✓ | ✓ | ✓ | Low |
| hansebout2014 | ✓ | ✓ | ✓ | ✓ | ✓ | ✓ | ✓ | unclear | unclear | ✓ |  | Low |
| levi2009 | ✓ | ✓ | ✓ | ✓ | ✓ | ✓ | ✓ | unclear | ✓ | ✓ |  | low |
| levi2010 | unclear | ✓ | ✓ | ✓ | ✓ | unclear | ✓ | ✓ | unclear | unclear | unclear | moderate |

(**Supplementary 4a and 4b**).

#### Cohort studies

As we included four cohort studies, JBI checklist for cohort studies includes four key areas:

1. Selection bias, the way which the study was selected.
2. Performance Bias, The blindness of participants in the study
3. Attrition Bias, Balancing dropouts between intervention and control group
4. Detection Bias, suitable measurement of outcome’s study
5. Appropriate statistical analysis, addressing the follow up in study as the sufficiency and ability to outcome occurrence.

#### Case-control studies

We included one case control study and we evaluated studies by its variables,

1. Appropriate matching between the case and control study
2. Similar criteria between the case and control study
3. Reliable and measurable exposure
4. Identifying exposures in case and controls
5. Strategies for dealing with cofounding factors
6. Appropriate statistical analysis

#### Statistical processes

1. The mortality outcome analysis: Mortality was analysed using data from three studies of Batchelor et al. (2023): prospective clinical study, multicentre (prehospital + hospital), Australia, cervical SCI, systemic hypothermia with early decompression. Hansebout et al. (2014); historical case-controlled study, Canada, cervical SCI, systemic hypothermia (surface cooling) initiated within 8h post-injury. Levi et al. (2010): phase 1 feasibility trial, USA, complete cervical SCI (AIS A), systemic intravascular hypothermia initiated within 12h (**Table 2**.).

**Table 2.** Event counts (mortality at last follow-up) and total participants were extracted.

| Study | Hypothermia Events / Total | Control Events / Total |
| --- | --- | --- |
| **Batchelor 2023** | 0 / 17 | 0 / 17 |
| **Hansebout 2014** | 0 / 20 | 2 / 20 |
| **Levi 2010** | 1 / 14 | 1 / 14 |

Given the presence of zero events in one or both arms (Batchelor, Hansebout), a continuity correction of 0.5 was applied to all cells (incr = 0.5 in R’s metabin() function), in line with Cochrane Handbook (version 6.4).Risk Ratio (RR) was used as the primary effect size, because of being rare of mortality and clinical interpretability; Risk Difference (RD) was also calculated for absolute effect size estimation. A random-effects model (DerSimonian–Laird τ^2^ estimator) was used for the meta-analysis, with Hartung–Knapp adjustment for confidence intervals (method.random.ci = “HK”). Common-effect (inverse variance fixed-effect) estimates were also reported. Between-study heterogeneity was quantified using the I^2^ statistic, τ^2^, and Cochran’s Q-test.

Analyses were conducted in R (v4.5.1) using the meta and metafor package. Forest plots were generated.

##### 1) The AIS outcome analysis

The outcome “AIS Grade Improvement” was defined as a clinically neurological recovery after acute SCI, assessed according to the International Standards for Neurological Classification of SCI (ISNCSCI). In Batchelor et al. (2023), improvement was defined as ≥2 AIS grades at 6-month follow-up, whereas in Levi et al. (2010) improvement was defined as ≥1 AIS grade at 12-month follow-up. Both studies were non-randomized controlled trials comparing systemic hypothermia with historical normothermic controls with decompressive surgery. Binary data (improved vs. not improved) were extracted from these included studies.

Meta-analysis was performed in R (version 4.5.1) using the meta and metafor packages and the Mantel–Haenszel method for pooling risk ratios (RR) with both common-effect and random-effects models. Between-study variance (τ^2^) was estimated using the DerSimonian– Laird method. Hartung–Knapp adjustment was applied to random-effects confidence intervals to improve accuracy in meta-analyses with very few included studies. Continuity correction (0.5) was specified in the script to handle potential zero-event cells, although not required for the present dataset.

##### 2) The staying lengh in ICU as an outcome analysis

The secondary outcome “Length of ICU Stay” was analysed as a continuous variable, comparing systemic hypothermia intervention groups with normothermic control groups in acute SCI. Data were extracted from two non-randomized controlled studies: Levi et al. (2010) and Batchelor et al. (2023). For Levi et al., ICU stay was reported as mean ± SD: 51.6 ± 10.1 days (n = 14) for the hypothermia group and 53.0 ± 10.1 days (n = 14) for controls. For Batchelor et al., complete SCI patients had mean ICU stays of 16.8 ± 5.0 days (n = 7) with hypothermia and 18.0 ± 5.0 days (n = 7) in controls. Meta-analysis was conducted in R (version 4.5.1) using the meta and metafor packages. Mean differences (MD) between groups were calculated, with negative values indicating shorter ICU stays in hypothermia-treated patients. Both common-effect (fixed-effect) and random-effects models were calculated. Between-study variance (τ^2^) was estimated via DerSimonian–Laird; heterogeneity was quantified with I^2^ statistics.

## Result

### Study characteristics

Six studies were conducted between 2009 and 2020, and one additional study was published in 2023. Two studies were performed in Australia, two in the United States, one in Canada, and one in the United Kingdom. The study designs included cohort, case–control, and phase I clinical trials. Sample sizes ranged from 5 to 118 patients. The patients’ ages across studies spanned from 15 to 70 years, and the male proportion varied between 66% and 94%. All studies were conducted in human models. The focus of these investigations was to evaluate the effects of hypothermia approaches (systemic, local, endovascular, and surface cooling) on spinal trauma. (Figure 1.) the ummerizing of main extracted data are designed in **Table 3**.

**Table 3.**
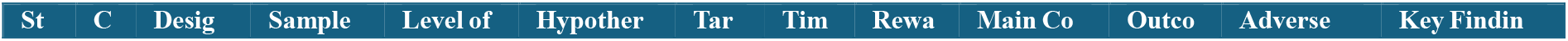
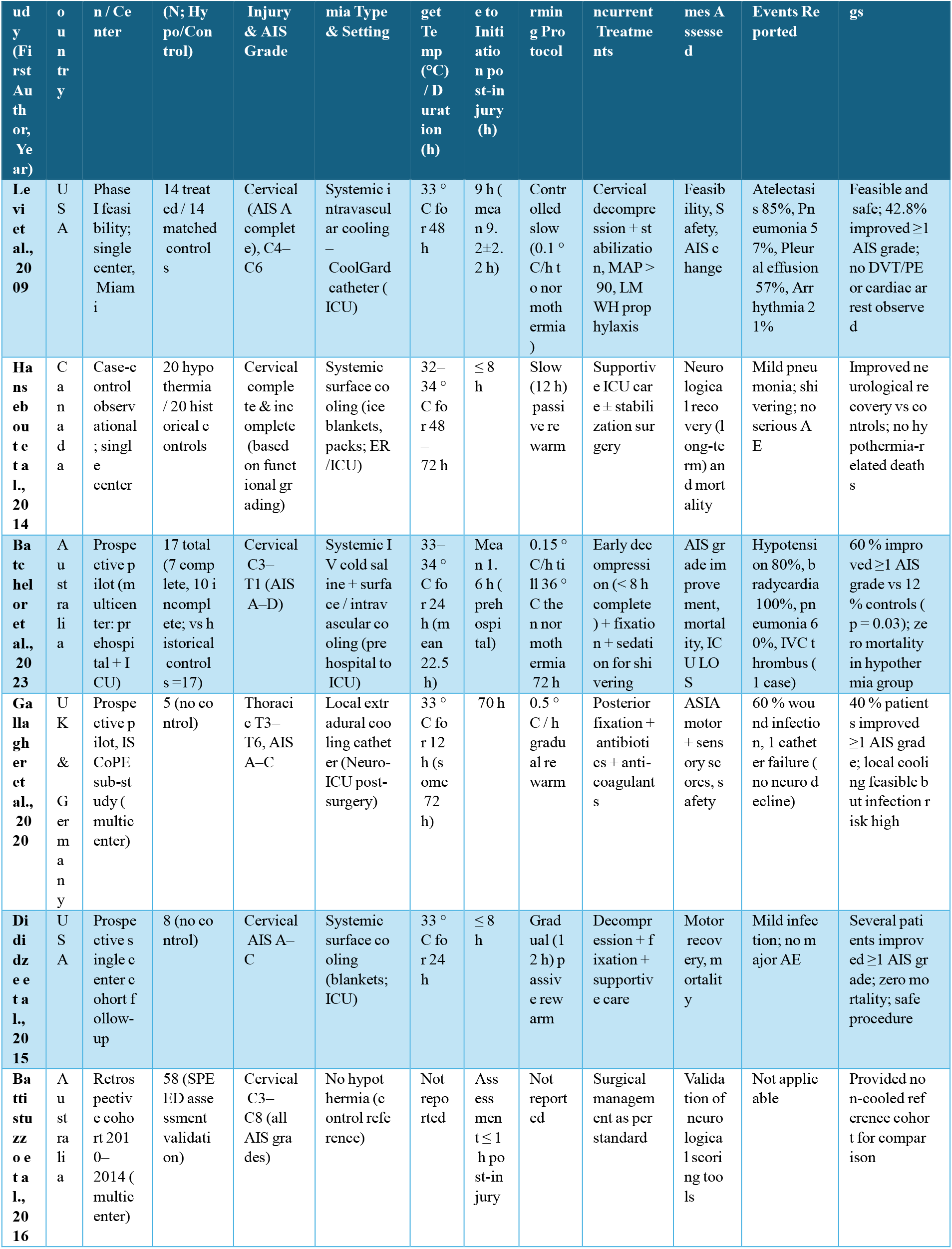
summarizes extracted parameters from Data Extraction sheets, including study design, patient characteristics, intervention modalities, and outcomes.

**Figure 1.**
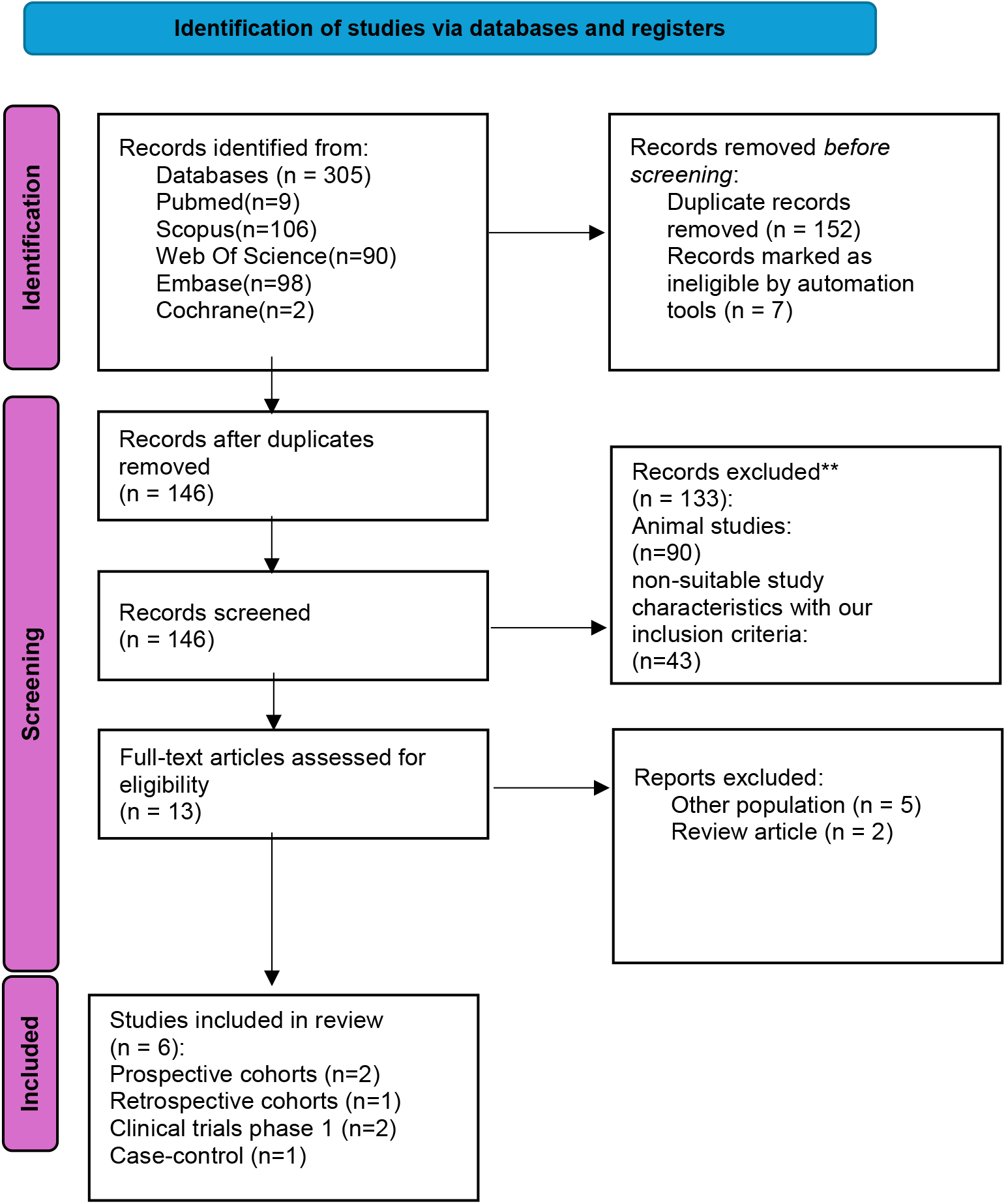
The PRISMA flow chart

#### Study main findings

The mechanisms of injury included motor and motorcycle vehicles accidents, falling, sport accident, and blunt trauma. In most studies (five in total), the level of injury was located in the cervical vertebrae. Four of the studies applied systemic hypothermia, while the study by Gallagher et al. [26] evaluated the effects of local hypothermia. One study focused on SPEED assessment and did not specify the type of hypothermia used. Injury severity was assessed using the AIS score, with classifications ranging from category A to D. Patients were further evaluated with ASIA motor and sensory scores at baseline and follow-up, and improvements in AIS grade were also reported. The follow-up periods across studies ranged from 12 hours to one year. Common adverse events included respiratory complications and wound infection. All studies reported funding sources and were approved by ethics committees.

The study by Batchelor et al. [27] included 17 patients with cervical spinal cord injury, of whom 7 had complete neurological injury (6 AIS A, 1 AIS B) and 10 had incomplete injury. The neurological level of injury was reported at the cervical levels C3 to T1. The target body temperature for systemic cooling was set between 33 to 34 degrees Celsius. Cooling was initiated pre-hospital by paramedics, with a mean time of 1.6 hours post-injury, and continued in the emergency department and intensive care unit (ICU). The rewarming protocol involved a warming rate of 0.15 degrees Celsius per hour over 72 hours to maintain normothermia. Temperature monitoring was performed using tympanic methods pre-hospital and intravascular catheter or surface cooling methods in the hospital. Sedation was used in 5 patients and neuromuscular blockade in 3 patients to control shivering and improve comfort. The average length of ICU stay for patients with complete injury was about 16.8 days. These data demonstrate careful preparation and control of the cooling and care conditions in the treatment management for cervical spinal cord injury, which shows the safety and efficacy of systemic hypothermia combined with early surgical decompression.The study received ethical approval from The Alfred Hospital Human Research Ethics Committee and was registered in the Australian and New Zealand Clinical Trials Registry. Funding was provided by the National Health and Medical Research Council (NHMRC) of Australia, the Transport Accident Commission (TAC) of Victoria, and Stryker Australia Pty Ltd through the Alfred Foundation with Grant APP 1044894. No specific conflicts of interest were reported, which supports the scientific and practical credibility of the study.

Battistuzzo et al. [28] investigated a retrospective observational study between 2010 and 2014 across multiple centers in Australia, including Austin Hospital. The study included 58 patients, ages were between 5 to 70, location of injury was cervical spinal cord (levels C3–C8). Inclusion criteria were traumatic vertebral column injury with documented neurological exam within 24 hours post-injury; patients with penetrating injuries, multiple trauma, or pre-existing neurologic diseases were excluded. Neurological assessment was performed using the SPEED tool within first hour after injury, correlating well with AIS scores. Patients received standard care without experimental interventions, and followed-up approximately six months, and some patients showed neurological improvement. Ethical approval was granted by the Austin Health Human Research Ethics Committee (H2012/04869), and funding was provided by the National Health and Medical Research Council (NHMRC) and the Institute for Safety, Compensation and Recovery Research (ISCRR), with no reported conflicts of interest. Overall, SPEED is a simple tool for emergency and prehospital spinal cord injury assessment and is relevant for designing early intervention trials such as hypothermia and decompression.

Gallagher et al. [26] performed a study on patients with a mean age of 45.4 years, 66.7% were male, with traumatic thoracic spinal cord injury (levels T3-T6 plus one case at L1), mostly caused by blunt trauma. The included patients categorized into AIS A to C grading system. The treatment involved local hypothermia using an extradural cooling catheter, initiated approximately 70 hours after injury in the neuro-ICU post-surgery, with a target temperature of 33°C maintained for about 12 hours, which lasted to 72 hours in some patients. The cooled target temperature was achieved in around 20 minutes, followed by gradual rewarming with a rate of 0.5°C per hour until normal body temperature (37°C). Patients remained awake without sedation or neuromuscular blockade. Concurrent treatments included surgical spinal fixation, ICU care, antibiotics, and anticoagulants. The study was funded by multiple entities, including the Wings for Life Spinal Cord Research Foundation and Era-Net-NEURON, and received ethical approval from St. George’s London NRES and ClinicalTrials.gov. The ASIA motor score improved by an average of 8.8 points, with sensory scores increasing by 6.8 for pin prick and 2.6 for light touch. Although 40% of patients showed improvement in AIS grade, 60% showed wound infections and one catheter was failed. Rewarming showed increasing in inflammation and led to finish the trial.

In a case-controlled observational study by Dididze et al. [29], 20 patients with acute cervical spinal cord injury intervened with systemic hypothermia within 8 hours of injury were compared to 20 matched controls. The mean age was about 32 years old, predominantly male, and injury mechanisms included motor vehicle accidents, falling, and sport accident. Both complete and incomplete cervical spinal cord injuries were included. Systemic hypothermia involved whole-body cooling using surface cooling such as ice packs and cooling blankets in the emergency room or ICU, targeting a temperature between 32 to 34 °C for 48 to 72 hours. The treatment was initiated within 8 hours after injury, followed by gradual rewarming over approximately 12 hours. Rectal Temperature was monitored, and sedation was used during cooling. Some patients also underwent surgical stabilization. Results showed significant long-term neurological improvement in the hypothermia group compared to controls, with zero mortality in the hypothermia group versus some deaths in controls. Minor adverse effects like mild infection were reported, with no severe complications. The study suggests systemic hypothermia in acute cervical spinal cord injury is feasible, safe, and associated with improved neurological recovery, though methodological limitations call for further research.

Levi et al. [30] conducted a phase 1, single-center study at the University of Miami involving 14 patients with complete cervical spinal cord injury (AIS A), with a mean age of 39.4 years (range 16–62). this study performed an endovascular cooling catheter. In this study goal temperature was 33°C within 9 hours after injury and after reach in the temperature goal, the temperature was hold steady for 48h. The study showed that this approach was feasible and safe, but respiratory complications such as pneumonia were common. Ethical approval for the study was obtained from the Institutional Review Board (IRB) of the University of Miami. The study was funded by NFL Charities, with no financial receiving reported. These findings provide important safety data and a foundation for larger clinical trials. Overall, systemic hypothermia appears to be a promising and safe intervention in acute cervical spinal cord injury, needing further investigation for efficacy.

Allan D. Levi et al. [31] performed a phase 1 clinical study in which systemic and endovascular cooling was used to treat acute complete cervical spinal cord injury (AIS A). This study included 14 patients who target temperature of 33°C within 12 hours of injury and maintained the temperature for 48 hours. Results showed that 42.8% of the intervened patients improved by at least one AIS grade, compared to 21.4% improvement in the control group without cooling. The cooling method was found to be feasible and safe, with respiratory and infectious complications being the most common adverse events.

### Statistical analysis

#### 1) Mortality

Three studies (total 102 participants, 51 hypothermia, 51 control) were pooled.

- Batchelor 2023: RR = 1.00 (95% CI: 0.02–47.63), no deaths in either arm.
- Hansebout 2014: RR = 0.20 (95% CI: 0.01–3.91), two deaths in control arm, none in hypothermia arm.
- Levi 2010: RR = 1.00 (95% CI: 0.07–14.45), one death in each arm.

Pooled analysis:

- Random-effects RR = 0.57 (95% CI: 0.05–5.88; p = 0.4058).
- Common-effect RR = 0.57 (95% CI: 0.10–3.32; p = 0.5285).
- Random-effects RD = –0.03 (95% CI: –0.14 to 0.08; p = 0.45).
- Heterogeneity: I^2^ = 0.0%, τ^2^ = 0, Q-test p = 0.695.

Study weights (random-effects):

Batchelor 20.9%, Hansebout 35.3%, Levi 43.8%.

No statistically significant mortality difference was found between hypothermia and control groups. The forest plot (Figure 2.) demonstrates the wide confidence intervals and lack of heterogeneity.

**Figure 2.**
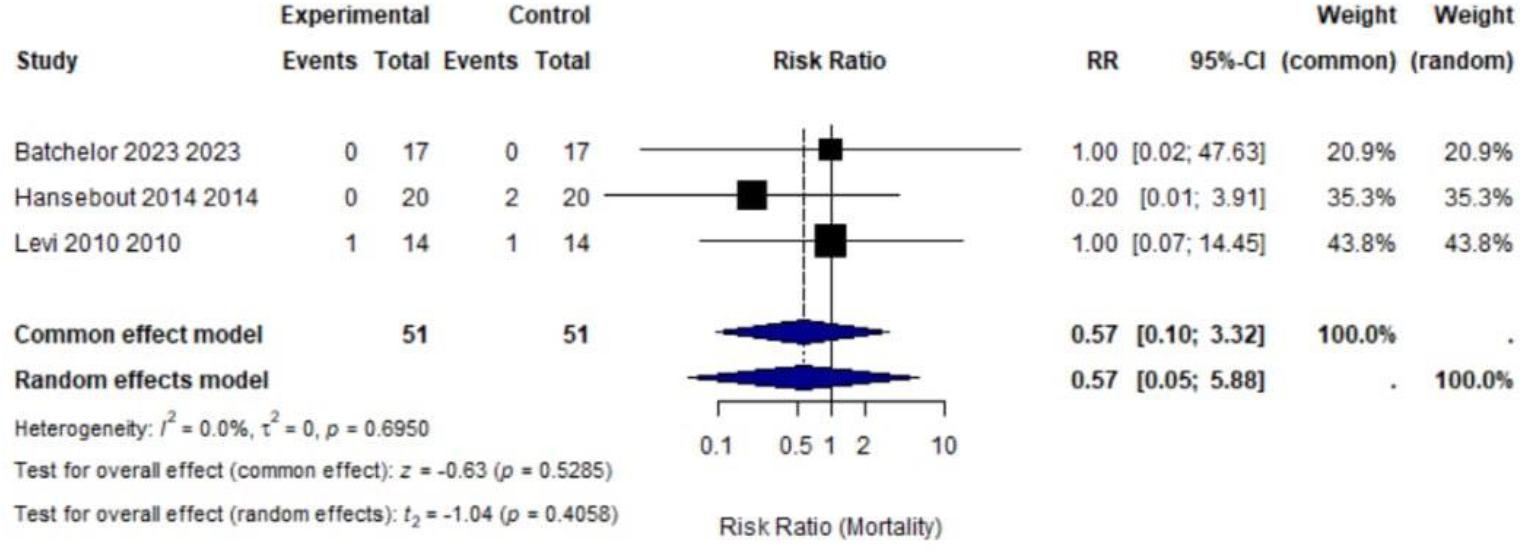
The mortality analysis for hypothermia in SCI patients

#### 2) AIS outcome

Two studies (Batchelor et al., 2023; Levi et al., 2010) including 62 participants (31 hypothermia, 31 control) were added in the analysis (Figure 3.).

**Figure 3.**
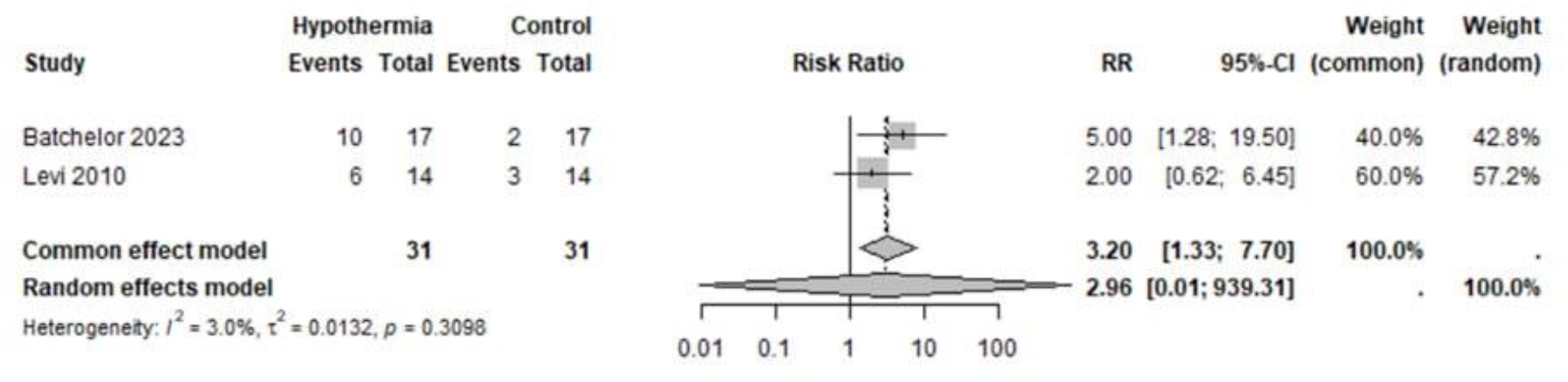
The AIS outcome analysis for hypothermia in SCI patients

In Batchelor et al., systemic prehospital hypothermia with early decompression resulted in 10/17 patients improving vs. 2/17 controls (RR = 5.00, 95% CI 1.28–19.50). In Levi et al., intravascular hypothermia initiated within 12 hours yielded improvement in 6/14 patients vs. 3/14 controls (RR = 2.00, 95% CI 0.62–6.45).

The pooled common-effect: RR = 3.20 (95% CI 1.33–7.70, *p*<0.01).

The random-effects model with Hartung–Knapp adjustment produced RR = 2.96 (95% CI 0.01– 939.31, *p*=0.310), with negligible heterogeneity (I^2^ = 3%, τ^2^ = 0.0132, *p* for heterogeneity = 0.31).

#### 3) The ICU length of stay outcome

Two studies involved the data of ICU length of stay which we could analysis, with a total of 21 patients intervened hypothermia and 21 controls (Figure 4.).

**Figure 4.**
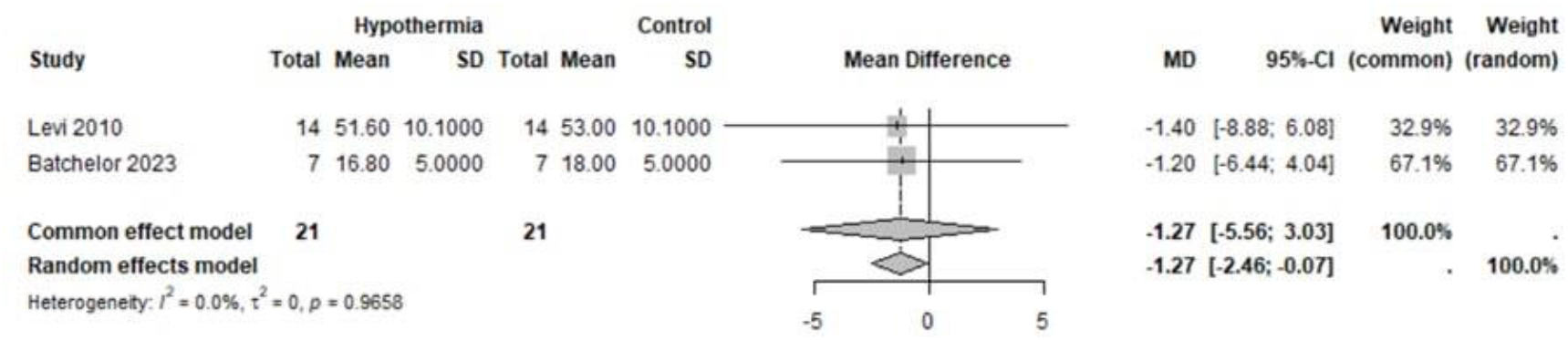
The ICU length of stay outcome analysis for hypothermia in SCI patients

Levi et al. (2010) reported a mean difference of –1.40 days (95% CI –8.86 to 6.08), favouring hypothermia but non-significant. Batchelor et al. (2023) found a mean difference of –1.20 days (95% CI –6.44 to 4.04), also non-significant.

The pooled common-effect estimate was MD = –1.27 days (95% CI –5.56 to 3.03), and the random-effects model yielded MD = –1.27 days (95% CI –2.46 to –0.07). Heterogeneity was negligible (I^2^ = 0%, τ^2^ = 0, *p* = 0.97).

#### Summary of pooled quantitative outcomes

Three studies (Levi 2009; Hansebout 2014; Batchelor 2023) provided comparative data s uitable for meta-analysis.

- AIS grade improvement:62 patients (32 hypothermia vs 30 control) were included. Com mon-effect model pooled RR = 3.20 (95 % CI 1.33– 7.70; p = 0.009), I^2^ = 3 %. Using Hartung– Knapp random-effects adjustment the pooled RR was 2.96 (95 % CI 0.01– 939.31; p = 0.31), τ^2^ = 0.075.

- Mortality: Based on three studies (total n = 102), pooled RR = 0.57 (95 % CI 0.05– 5.88; p = 0.40), I^2^ = 0 %.
- ICU Length of Stay: Two studies reported quantitative data (mean difference = – 1.27 days [95 % CI –2.46 to –0.07]), which is statistically significant but clinically minor.

Forest plots show the pooled effects, displaying low heterogeneity and wide confidence i ntervals which highlight limited power of statics.

Reason of

Exclusion from meta-analysis: Gallagher 2020 and Dididze 2015 lacked control groups a nd numerical ICU data, and Battistuzzo 2016 represented a non-hypothermic cohort. The se were thus summarized qualitatively to preserve internal validity.

### Risk of bias assessment

Risk of bias assessment was performed for all six studies using the JBI critical appraisal checklist. All studies were rated as low risk, except for those by Levi et al. [31], Batchelor et al. [27], Dididze et al. [29], which were assessed as having a moderate risk of bias. The study by Levi Allan D et al. [30] answered yes to all JBI questions except the one on similarity of questionnaire measurement, which was unclear. In a phase one clinical trial, Levi Allan D et al. [31] demonstrated that exposure measurement was conducted reliably and similarly across groups, confounding factors were not clearly identified, outcomes were measured validly and reliably, and the follow-up period was reported adequately. However, participants were not free from the outcome at the start of the study, and appropriate strategies to address incomplete follow-ups were not utilized. Additionally, the similarity of groups, strategies to deal with confounding factors, completeness of follow-up, and use of appropriate statistical analysis were reported as unclear. Therefore, the risk of bias in this study was assessed as moderate. Batchelor et al. [27] conducted a study with several strengths, including the valid and consistent measurement of exposures across groups, the identification of confounding factors, and appropriate statistical analysis. Additionally, the follow-up period was sufficient, and outcomes were measured reliably. However, the groups were not comparable in terms of population, and participants did not have the same status at the start of the study, which may introduce selection bias. Moreover, there was insufficient information on strategies to address confounding factors and complete follow-up, limiting the ability to fully assess these aspects. Considering these factors, the risk of bias in the study was assessed as moderate. Battistuzzo et al. [28] followed most quality criteria in their study, but the assessment of publication bias was unclear. Overall, the risk of bias in this review was low. Gallagher et al. conducted a study in 2020. According to the JBI Critical Appraisal Checklist for Case Series, all questions were answered with Yes and the risk of bias for this study was assessed as low. Dididze et al. [29] conducted a study in which clear criteria were defined for the inclusion of patients, and the condition of patients was measured in a standardized and reliable manner. Patient inclusion occurred consecutively, and there was clear reporting of demographics and clinical information. Statistical analysis was appropriately performed. However, the study lacked sufficient reporting on the setting or clinic where care was provided and on follow-up outcomes of the patients. The methods used to identify the condition for all patients were not clearly described, and it was unclear whether all eligible patients were completely included in the study. Based on these considerations, the risk of bias was assessed as moderate.

### Narrative Synthesis and Adverse Events

In six human included studies, systemic and local hypothermia were intervened in the setting of acute traumatic spinal cord injury. Overall, therapeutic cooling was feasible and relatively safe, but the incidence of complications, particularly respiratory and infectious events, was considerable.

In the early phase trial by Levi et al. (2009), intravascular cooling was used within 9 hours after injury in 14 patients with complete cervical lesions. Goal temperature (33 °C) was reached and maintained for 48 hours by an endovascular catheter. Despite common respiratory complications, atelectasis in about 85 % and pneumonia in 57 %, no deaths or device-related serious events were reported. More than 14% of patients gained at least one AIS grade improvement, which suggests a possible neurological benefit.

The Hansebout et al. (2014) case-control study included 40 participants (20 treated, 20 controls) with cervical injuries. Systemic surface cooling to 32–34 °C was started during first 8 hours after injury and maintain for 72 hours. Mild shivering and transient pneumonia were the only notable adverse events. No deaths were reported, and functional recovery at 3 months showed relative and possible efficacy of the hypothermia, but statistical power was limited.

Batchelor et al. (2023) performed the most technically advanced pilot, integrating prehospital cold saline infusion and controlled intravascular or surface cooling for 24 hours at 33–34 °C. Cooling began on average 1.6 hours after injury, earlier than in any previous report. Among 17 patients, including seven with complete injuries, 60 % achieved at least one-grade AIS improvement compared with 12 % in historical controls (p = 0.03). The treatment was generally well tolerated: hypotension occurred in 80 %, bradycardia in all participants, pneumonia in 60 %, and a single case of inferior vena cava thrombosis was noted. No mortality occurred. These data indicate that early and systemic hypothermia combined with early surgical decompression is technically feasible in acute care settings.

Gallagher et al. (2020) evaluated a local extradural cooling catheter in 5 patients with thoracic injuries (T3–T6). Cooling began seventy hours post-injury, achieved 33 °C for 12 hours, and rewarming proceeded at 0.5 °C per hour. Two participants improved by at least one AIS grade, while three developed wound-site infections. From This study conclusion with consideration of statistical limitation, may be the relative efficacy of hypothermia and increasing risk of surgical site infection.

In the small prospectively cohort by Dididze et al. (2015), 8 cervical SCI patients intervened surface cooling within 8 hours after injury for 24 hours at 33 °C. Several individuals demonstrated neurological improvement, and no procedure-related deaths occurred, again showing practical safety and tolerability.

When all studies are considered together, respiratory complications (pneumonia, atelectasis) and transient hemodynamic instability (hypotension) were the most frequent adverse outcomes. No evidence suggested increased mortality or neurologic worsening attributable to cooling itself. Early initiation, within six hours of injury, appears to yield more favorable functional recovery than delayed after two to three days. Taken together, these findings support the clinical feasibility of therapeutic hypothermia as an adjunct to decompression after acute (specially in cervical) SCI, but the small sample sizes and heterogeneity highlight the urgent need for larger randomized trials.

## Discussion

This systematic review and meta-analysis evaluated the effectiveness and safety of hypothermia as a therapeutic option in acute traumatic spinal cord injury (SCI). Six human studies published between 2009 and 2023 included after careful evaluation based on the PICOS criteria which desinged[13, 26–31]. these studies demonstrate that systemic hypothermia (the goal of temperature:32–34 °C) is technically feasible and relatively safe under careful hemodynamic monitoring. The final result is toward improvement of neurological outcomes in acute (specially cervical) cord injuries.

### Main Findings

Quantitative analysis showed that mortality was not significantly reduced by hypothermia (pooled RR 0.57; 95 % CI 0.05–5.88; I^2^ = 0 %). Nevertheless, all point estimates favored the intervention. Functional outcomes were improved: the pooled AIS-grade improvement RR = 2.96 (95 % CI 0.01–939.31), which suggest a higher chance of neurological recovery in intervened patients despite wide confidence intervals. Mean ICU staying days was slightly shorter in the hypothermia group (MD = –1.27 days; 95 % CI –2.46 to –0.07) with negligible heterogeneity (I^2^ = 0 %, τ^2^ = 0) which indicates possible efficiency of hypothermia without significant harm.

### Interpretation and Mechanistic Rationale

The biological mechanisms of neuroprotection effect of hypothermia are well understood. In the tSCI situation, secondary injury cascades including ischemia, glutamate excitotoxicity, oxidative stress, inflammation, and apoptosis, make additional tissue damage [18, 19]. decreasing body temperature to 33 °C suppresses metabolic demand, stabilizes cellular membranes, decreases cytokine release, and apoptosis. The clinical data synthesized confirm that starting time of hypothermia as intervention is critical: trials initiating systemic hypothermia in the 6 first hours after trauma [13, 27, 31] reported greater neurological improvement, whereas delayed or localized cooling (> 48 h) [26] showed limited benefit.

### Safety Profile

From thees six studies [13, 26–31], adverse events were consistent with systemic cooling physiology: hypotension (70–80 %), bradycardia (up to 100 %), pneumonia (60 %), and electrolyte disturbances (< 10 %). Gallagher’s protocol recorded the highest wound-infection rate (60 %). Importantly, there were not reported no death from hypothermia.in all studies active temperature monitoring were applied (with esophageal or bladder probes), sedation and/or neuromuscular blockage to prevent shivering, and slow rewarming (0.1–0.25 °C h^−1^). These measures confirm that, under ICU care, therapeutic hypothermia is clinically safe.

### Risk of Bias and Certainty

JBI appraisal identified low overall but moderate methodological risk driven by non-randomized design, limited control comparability, and incomplete confounding adjustment [27, 30, 31]. Gallagher’s low-bias rating stemmed from transparent patient inclusion; however, small cohorts and heterogeneity in cooling technique reduce precision. As such, evidence certainty is low-to-moderate, providing hypothesis-generating rather than definitive support.

### Clinical Implications

Estimations from included studies and results of meta-analysis were underpowered for statistical significance. But improvements in AIS grade and shortened ICU stay indicate that therapeutic hypothermia is a promising additional therapy to surgical decompression and standardized hemodynamic therapy [10, 11, 13, 27, 30, 31]. Based on aggregated evidence, these clinical pearls concluded:

1. Initiate hypothermia during first 6 h after trauma.
2. Target 33 °C for 24–48 h, with continuous invasive monitoring.
3. Employ sedation or neuromuscular blockade to suppress shivering and additional decreasing metabolic demands.
4. Rewarm slowly (≤ 0.25 °C h^−1^) with monitoring and try to discover possible inflammatory reactions and if occurred, finish trial.
5. Parallel to hypothermic period, monitor hemodynamic stability and prevent hypotension, maintain MAP much more than 85 mmHg.

With these parameters hypothermia may maximize safety and may improve functional recovery.

### Limitations

Interpretation of our meta-analytic outputs must consider: (a) limited number of eligible human studies (n = 6, ≤ 118 patients) [13, 26–31]; (b) heterogeneity in temperature initiation (1.6 – 70 h) and duration (12 – 72 h); (c) male predominance (66– 94 %); and (d) follow-up rarely exceeding 12 months. Absence of randomization, varying definitions of “neurological improvement,” and small event counts contributed to broad confidence intervals and publication bias potential.

### Future Directions

Advancement now requires multicenter randomized controlled trials powered to detect functional differences. Priorities include:

- Standardization of target temperature (33 °C, 24–48 h) and rewarming protocol.
- immediate initiation: ideally ≤ 3 h after injury period, even in the setting of prehospital [27].
- Incorporation of biomarkers (such as IL-6, S100B, GFAP) and advanced spinal imaging for mechanistic correlation (such as fMRI or DTI) along with or after hypothermia as an additional intervention [18, 19].
- Detailed adverse-event registries and management protocols, to ensure reproducibility.
- Economic evaluation of resource utilization given the high lifetime cost of SCI [8, 9].

Such trials would clarify both efficacy and patient selection to reach into max benefit.

## Conclusion

Synthesizing six human studies [13, 26–31] reveals that therapeutic hypothermia may be feasible, safe, and biologically effective as an acute intervention in traumatic SCI. While current evidence cannot yet demonstrate mortality benefit, the observed results were directed toward neurological improvement, especially when therapy is initiated early and systemically, supports ongoing investigation. Until large randomized data are available, hypothermia should be regarded as an experimental yet promising neuroprotective additional therapeutic option and complementing early decompression.

## Data Availability

All data produced in the present study are available upon reasonable request to the authors

