## Supplementory for "Safety and Efficacy of Therapeutic Hypothermia in Acute Traumatic Spinal Cord Injury: A Systematic Review and Meta-Analysis of human evidence-based studies"

**Search Strategy (n=305):**

**Scopus (n=106):**

TITLE-ABS-KEY ( "Spinal Cord Injury" OR "SCI" OR "acute spinal cord trauma" OR "spinal cord trauma" )
AND
(
TITLE-ABS-KEY ( "Hypothermia, Induced" OR "therapeutic hypothermia" OR "systemic hypothermia" OR "local hypothermia" OR "cooling therapy" OR "targeted temperature management" OR "regional hypothermia" )
OR
INDEXTERMS ( "hypothermia" )
)
AND
(
TITLE-ABS-KEY ( "Recovery of Function" OR "neurological recovery" OR "motor recovery" OR "sensory recovery" OR "neuroprotection" OR "secondary injury" OR "inflammation" OR "decreasing second insult" OR "anti inflammatory" )
OR
INDEXTERMS ( "recovery" OR "neuroprotection" )
)
AND
(
TITLE-ABS-KEY ( "acute" OR "early phase" OR "post-injury" OR "subacute" )
)

**PubMed (n=9):**

(("Spinal Cord Injury"[Mesh] OR "Spinal Cord Injuries"[Mesh] OR "SCI"[Title/Abstract] OR "acute spinal cord trauma"[Title/Abstract] OR "spinal cord trauma"[Title/Abstract])
AND
("Hypothermia, Induced"[Mesh] OR "therapeutic hypothermia"[Title/Abstract] OR "systemic hypothermia"[Title/Abstract] OR "local hypothermia"[Title/Abstract] OR "cooling therapy"[Title/Abstract] OR "targeted temperature management"[Title/Abstract] OR "regional hypothermia"[Title/Abstract] OR "hypothermia"[Mesh])
AND
("Recovery of Function"[Mesh] OR "neurological recovery"[Title/Abstract] OR "motor recovery"[Title/Abstract] OR "sensory recovery"[Title/Abstract] OR "neuroprotection"[Title/Abstract] OR "secondary injury"[Title/Abstract] OR "inflammation"[Title/Abstract] OR "decreasing second insult"[Title/Abstract] OR "anti inflammatory"[Title/Abstract] OR "recovery"[Mesh] OR "neuroprotection"[Mesh])
AND
("acute"[Title/Abstract] OR "early phase"[Title/Abstract] OR "post-injury"[Title/Abstract] OR "subacute"[Title/Abstract]))

**WOS (n=90):**

TS=(("Spinal Cord Injury" OR "Spinal Cord Injuries" OR "SCI" OR "acute spinal cord trauma" OR "spinal cord trauma")
AND
("Hypothermia, Induced" OR "therapeutic hypothermia" OR "systemic hypothermia" OR "local hypothermia" OR "cooling therapy" OR "targeted temperature management" OR "regional hypothermia" OR "hypothermia")
AND
("Recovery of Function" OR "neurological recovery" OR "motor recovery" OR "sensory recovery" OR "neuroprotection" OR "secondary injury" OR "inflammation" OR "decreasing second insult" OR "anti inflammatory" OR "recovery")
AND
("acute" OR "early phase" OR "post-injury" OR "subacute"))

**Embase (n=98):**

('spinal cord injury'/exp OR 'spinal cord injury':ti,ab OR 'spinal cord injuries':ti,ab OR 'sci':ti,ab OR 'acute spinal cord trauma':ti,ab OR 'spinal cord trauma':ti,ab)
AND
('induced hypothermia'/exp OR 'therapeutic hypothermia':ti,ab OR 'systemic hypothermia':ti,ab OR 'local hypothermia':ti,ab OR 'cooling therapy':ti,ab OR 'targeted temperature management':ti,ab OR 'regional hypothermia':ti,ab OR 'hypothermia'/exp)
AND
('recovery of function'/exp OR 'neurological recovery':ti,ab OR 'motor recovery':ti,ab OR 'sensory recovery':ti,ab OR 'neuroprotection':ti,ab OR 'secondary injury':ti,ab OR 'inflammation':ti,ab OR 'decreasing second insult':ti,ab OR 'anti inflammatory':ti,ab OR 'recovery'/exp OR 'neuroprotection'/exp)
AND
('acute':ti,ab OR 'early phase':ti,ab OR 'post-injury':ti,ab OR 'subacute':ti,ab)

**Cochrane (n=2):**

(
("Spinal Cord Injury" OR "Spinal Cord Injuries" OR "SCI" OR "acute spinal cord trauma" OR "spinal cord trauma")
AND
("Hypothermia, Induced" OR "therapeutic hypothermia" OR "systemic hypothermia" OR "local hypothermia" OR "cooling therapy" OR "targeted temperature management" OR "regional hypothermia" OR "hypothermia")
AND
("Recovery of Function" OR "neurological recovery" OR "motor recovery" OR "sensory recovery" OR "neuroprotection" OR "secondary injury" OR "inflammation" OR "decreasing second insult" OR "anti inflammatory" OR "recovery")
AND
("acute" OR "early phase" OR "post-injury" OR "subacute")
)
