## Supplementory for "Safety and Efficacy of Therapeutic Hypothermia in Acute Traumatic Spinal Cord Injury: A Systematic Review and Meta-Analysis of human evidence-based studies"

1) JBI Critical Appraisal Checklist for Case Series :**clinical application of modest hypothermia after spinsl cord injury**

Reviewe	مبینا قمرپورDate20/6/1404 یا 11/9/2025

Author:Levi.AllanD	Year2009	Record Number:10.1089/neu.2008.0745

|  | Yes | No | Unclear | Not applicable |
| --- | --- | --- | --- | --- |
| Were there clear criteria for inclusion in the case series? | □ 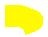 | □ | □ | □ |
| Was the condition measured in a standard, reliable way for all participants included in the case series? | □ 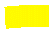 | □ | □ | □ |
| Were valid methods used for identification of the condition for all participants included in the case series? | □ 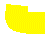 | □ | □ | □ |
| Did the case series have consecutive inclusion of participants? | □ 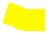 | □ | □ | □ |
| Did the case series have complete inclusion of participants? | □ 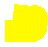 | □ | □ | □ |
| Was there clear reporting of the demographics of the participants in the study? | □ 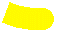 | □ | □ | □ |
| Was there clear reporting of clinical information of the participants? | □ 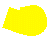 | □ | □ | □ |
| Were the outcomes or follow up results of cases clearly reported? | □ | □ | □ 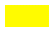 | □ |
| Was there clear reporting of the presenting site(s)/clinic(s) demographic information? | □ 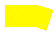 | □ | □ | □ |
| Was statistical analysis appropriate? | □ 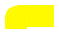 | □ | □ | □ |

2)JBI Critical Appraisal Checklist for cohort studies**: clinical outcomes using modest intravascular hypothermia after acute cervical spinal cord injury**

Review: **Mobina ghamarpour** _ Date:20/6/1404

Author:**Levi.Allan.D/** Year:**2010/** Record Number:**10.1227/01.NEU.0000367557.77973.5F**

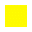

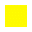

|  | Yes | No | Unclear | Not applicable |
| --- | --- | --- | --- | --- |
| 1. Were the two groups similar and recruited from the same population? | □ | □ | □ 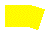 | □ |
| 2. Were the exposures measured similarly to assign people to both exposed and unexposed groups? | □ 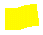 | □ | □ | □ |
| 3. Was the exposure measured in a valid and reliable way? | □ 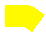 | □ | □ | □ |
| 4. Were confounding factors identified? | □ 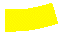 | □ | □ | □ |
| 5. Were strategies to deal with confounding factors stated? | □ | □ | □ 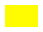 | □ |
| 6. Were the groups/participants free of the outcome at the start of the study (or at the moment of exposure)?  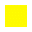 | □ | □ 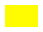 | □ | □ |
| 7. Were the outcomes measured in a valid and reliable way? | □ 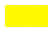 | □ | □ | □ |
| 8. Was the follow up time reported and sufficient to be long enough for outcomes to occur? | □ 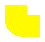 | □ | □ | □ |
| 9. Was follow up complete, and if not, were the reasons to loss to follow up described and explored? | □ | □ | □ 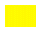 | □ |
| 10. Were strategies to address incomplete follow up utilized? | □ | □ 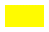 | □ | □ |
| 11. Was appropriate statistical analysis used? | □ | □ | □ 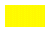 | □ |

3)JBI Critical Appraisal Checklist for cohort studies :immediate cooling and early decompression for the treatment of cervical spinal cord injury

Review: **Mobina ghamarpour** Date20**/6/1404**

Author:**Batchelor.PE/** Year:**2023/** Record Number: 10.1089/ther.2022.0046

|  | Yes | No | Unclear | Not applicable |
| --- | --- | --- | --- | --- |
| 1. Were the two groups similar and recruited from the same population? | □ | □ 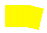 | □ | □ |
| 2. Were the exposures measured similarly to assign people to both exposed and unexposed groups? | □ 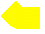 | □ | □ | □ |
| 3. Was the exposure measured in a valid and reliable way? | □ 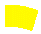 | □ | □ | □ |
| 4. 4Were confounding factors identified? | □ 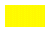 | □ | □ | □ |
| 5. Were strategies to deal with confounding factors stated? | □ | □ | □ 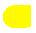 | □ |
| 6. Were the groups/participants free of the outcome at the start of the study (or at the moment of exposure)? | □ | □ 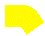 | □ | □ |
| 7. Were the outcomes measured in a valid and reliable way? | □ 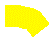 | □ | □ | □ |
| 8. Was the follow up time reported and sufficient to be long enough for outcomes to occur? | □ 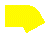 | □ | □ | □ |
| 9. Was follow up complete, and if not, were the reasons to loss to follow up described and explored? | □ | □ | □ 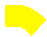 | □ |
| 10. Were strategies to address incomplete follow up utilized? | □ | □ | □ 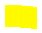 | □ |
| 11. Was appropriate statistical analysis used? | □ 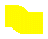 | □ | □ | □ |

4)JBI Critical Appraisal Checklist for systematic reviews and research syntheses studies: **EARLY RAPID NEUROLOGICAL ASSESSMENT FOR ACUTE SPINAL CORD INJURY TRIALS**

Reviewe: **Mobina ghamarpour /**Date:**1404/6/21**

Author:**caroline R.Battistuzzo** / Year: **2016** / Record Number: **10.1089/neu.2015.4360**

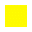

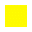

|  | Yes | No | Unclear | Not applicable |
| --- | --- | --- | --- | --- |
| 1. Is the review question clearly and explicitly stated? | □ 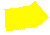 | □ | □ | □ |
| 2. Were the inclusion criteria appropriate for the review question? | □ 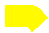 | □ | □ | □ |
| 3. Was the search strategy appropriate? | □ 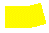 | □ | □ | □ |
| 4. Were the sources and resources used to search for studies adequate? | □ 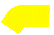 | □ | □ | □ |
| 5. Were the criteria for appraising studies appropriate? | □ 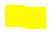 | □ | □ | □ |
| 6. Was critical appraisal conducted by two or more reviewers independently? | □ 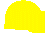 | □ | □ | □ |
| 7. Were there methods to minimize errors in data extraction? | □ 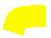 | □ | □ | □ |
| 8. Were the methods used to combine studies appropriate? | □ 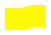 | □ | □ | □ |
| 9. Was the likelihood of publication bias assessed? | □ | □ | □ 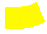 | □ |
| 10. Were recommendations for policy and/or practice supported by the reported data? | □ 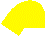 | □ | □ | □ |
| 11. Were the specific directives for new research appropriate? | □ 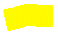 | □ | □ | □ |

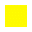

**5)JBI Critical Appraisal Checklist for Case Series :** **Effects of local hypothermia–rewarming on physiology, metabolism and inflammation of acutely injured human spinal cord**

Reviewer: **Mobina ghamarpour** /Date: **1404/6/21**

Author: **Mathew J. Gallagher** / Year:**2020**/ Record Number: **10.1038/s41598-020-64944-y**

|  | Yes | No | Unclear | Not applicable |
| --- | --- | --- | --- | --- |
| Were there clear criteria for inclusion in the case series? | □ 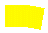 | □ | □ | □ |
| Was the condition measured in a standard, reliable way for all participants included in the case series? | □ 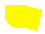 | □ | □ | □ |
| Were valid methods used for identification of the condition for all participants included in the case series? | □ 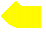 | □ | □ | □ |
| Did the case series have consecutive inclusion of participants? | □ 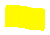 | □ | □ | □ |
| Did the case series have complete inclusion of participants? | □ 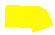 | □ | □ | □ |
| Was there clear reporting of the demographics of the participants in the study? | □ 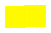 | □ | □ | □ |
| Was there clear reporting of clinical information of the participants? | □ 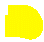 | □ | □ | □ |
| Were the outcomes or follow up results of cases clearly reported? | □ 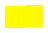 | □ | □ | □ |
| Was there clear reporting of the presenting site(s)/clinic(s) demographic information? | □ 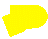 | □ | □ | □ |
| Was statistical analysis appropriate? | □ 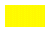 | □ | □ | □ |

6)JBI Critical Appraisal Checklist for Case Series : **Local cooling for traumatic spinal cord injury: outcomes in 20 patients and review of the literature**

Reviewer:**Mobina ghamarpour /** Date: **1404/6/21**

Author: **Robert R. Hansebout** / Year:**2014/**Record Number: **10.3171/2014.2.SPINE13318**

|  | Yes | No | Unclear | Not applicable |
| --- | --- | --- | --- | --- |
| Were there clear criteria for inclusion in the case series? | □ 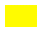 | □ | □ | □ |
| Was the condition measured in a standard, reliable way for all participants included in the case series? | □ 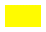 | □ | □ | □ |
| Were valid methods used for identification of the condition for all participants included in the case series? | □ | □ | □ 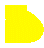 | □ |
| Did the case series have consecutive inclusion of participants? | □ 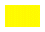 | □ | □ | □ |
| Did the case series have complete inclusion of participants? | □ | □ | □ 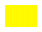 | □ |
| Was there clear reporting of the demographics of the participants in the study? | □ 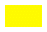 | □ | □ | □ |
| Was there clear reporting of clinical information of the participants? | □ 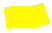 | □ | □ | □ |
| Were the outcomes or follow up results of cases clearly reported? | □ | □ 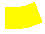 | □ | □ |
| Was there clear reporting of the presenting site(s)/clinic(s) demographic information? | □ | □ 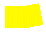 | □ | □ |
| Was statistical analysis appropriate? | □ 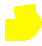 | □ | □ | □ |

| **author** | **title** | **Risk of bias** |
| --- | --- | --- |
| **Levi.AllanD** | clinical application of modest hypothermia after spinsl cord injury | **low** |
| **Levi.AllanD** | **clinical outcomes using modest intravascular hypothermia after acute cervical spinal cord injury** | **moderate** |
| **Batchelor.PE** | **immediate cooling and early decompression for the treatment of cervical spinal cord injury** | **moderate** |
| **caroline R.Battistuzzo** | **EARLY RAPID NEUROLOGICAL ASSESSMENT FOR ACUTE SPINAL CORD INJURY TRIALS** | **low** |
| **Mathew J. Gallagher** | **Effects of local hypothermia–rewarming on physiology, metabolism and inflammation of acutely injured human spinal cord** | **low** |
| **Robert R. Hansebout** | **Local cooling for traumatic spinal cord injury: outcomes in 20 patients and review of the literature** | **moderate** |
