## Supplementory for "Safety and Efficacy of Therapeutic Hypothermia in Acute Traumatic Spinal Cord Injury: A Systematic Review and Meta-Analysis of human evidence-based studies"

|  | Q1 | Q2 | Q3 | Q4 | Q5 | Q6 | Q7 | Q8 | Q9 | Q10 | Q11 | Overal |
| --- | --- | --- | --- | --- | --- | --- | --- | --- | --- | --- | --- | --- |
| Batchelor2023 | ⮾ | ☑ | ☑ | ☑ | ☑ | unclear | ☑ | ☑ | unclear | ⮾ | ☑ | moderate |
| battistuzzo2016 | ☑ | ☑ | ☑ | ☑ | ☑ | ☑ | ☑ | ☑ | ⮾ | ☑ | ☑ | Low |
| gallagher2020 | ☑ | ☑ | ☑ | ☑ | ☑ | ☑ | ☑ | ☑ | ☑ | ☑ | ☑ | Low |
| hansebout2014 | ☑ | ☑ | ☑ | ☑ | ☑ | ☑ | ☑ | unclear | unclear | ☑ |  | Low |
| levi2009 | ☑ | ☑ | ☑ | ☑ | ☑ | ☑ | ☑ | unclear | ☑ | ☑ |  | low |
| levi2010 | unclear | ☑ | ☑ | ☑ | ☑ | unclear | ☑ | ☑ | unclear | unclear | unclear | moderate |
